# Statistical fine-mapping of schizophrenia common risk loci using FINEMAP and SuSiE

**DOI:** 10.64898/2026.05.19.26353567

**Authors:** Eleanor R Mawson, Eilidh Fenner, James TR Walters, Michael C O’Donovan, Antonio F Pardiñas

## Abstract

The most recent Psychiatric Genomics Consortium (PGC) genome-wide association study of schizophrenia used the statistical fine-mapping tool FINEMAP to identify 70 genes that were likely to mediate common genetic variant associations with the disorder. Here, we extended that study by using two fine-mapping methods, SuSiE and FINEMAP, applying the methods to loci whose causal variant structure was considered too complex by the PGC, and optimising the proportion of posterior probability required by credible sets of causal SNPs for gene prioritisation. Prioritised gene sets were validated for schizophrenia relevance by testing for enrichment of loss-of-function mutation intolerance (LoFI), and for enrichment of rare deleterious coding variants associated with generalised cognition in UK Biobank, both known characteristics of schizophrenia associated genes. Concordance between FINEMAP and SuSiE was high, with most prioritised genes supported by both methods. Genes prioritised by both methods using a relaxed 80% posterior probability (PP) threshold for defining credible sets (N=98) were as enriched for LoFI and for rare deleterious missense variants associated with generalised cognition as genes prioritised using a more conservative 95% PP threshold (N=87). Loosening the credible set threshold combined with the joint application of SuSiE and FINEMAP increased the yield of prioritised genes by 40%, without reducing the orthogonal evidence for validity. Newly prioritised genes included calcium channel genes, *CACNA1I* and *CACNB2*, a glutamate receptor gene, *GRM3*, and *TCF4*, which has been previously implicated in schizophrenia.

## Introduction

Schizophrenia is an often highly debilitating disorder characterised by positive symptoms (e.g., hallucinations and delusions) and negative symptoms (e.g., apathy, social withdrawal, blunted affect). Globally, the disorder affects approximately 24 million people and ranks among the leading causes of disability^1^. Schizophrenia is highly heritable (twin-based h^2^ ∼80%)^2,3^ and polygenic^4^, with genetic risk conferred by both common and rare variation including copy number and ultra-rare coding variants^5^.

Genome-wide association studies (GWAS) have provided considerable insights into the common risk variant architecture of schizophrenia and other psychiatric disorders, but a key limitation is that the most significantly associated variant at a given locus is rarely the causal variant^6^. In order to make inferences from GWAS useful for downstream biological research, post-GWAS statistical fine-mapping is essential to identify the most likely underlying causal variants and, where possible, attribute them to specific genes.

The most recent Psychiatric Genomics Consortium (PGC) GWAS of schizophrenia compared 76,755 individuals with schizophrenia to 243,649 control individuals and found associations at 287 distinct genomic loci^7^. Loci were fine-mapped using FINEMAP^8^, and genes defined as “prioritised” if all credible causal SNPs within a locus were contained within a single gene, or if a SNP with a posterior probability (PP) greater than 0.1 was a non-synonymous or untranslated region (UTR) variant based on the Ensembl Variant Effect Predictor (VEP)^9^. This approach yielded 70 FINEMAP prioritised genes (64 protein-coding).

Three features of the PGC analysis limit its scope. First, gene prioritisation was limited to loci where FINEMAP predicted three or fewer causal variants, leaving complex loci uncharacterised. Second, at loci where FINEMAP (as well as conditional association analyses) suggested multiple causal variants, conservatively, all credible SNPs were required to map within the same gene, overlooking loci in which independent signals could point to different genes. Third, only one fine-mapping tool was applied, although alternatives based on different underlying methodologies exist, such as the Sum of Single Effects (SuSiE) model^10^.

Here we addressed these limitations, expanding on the work of the PGC, by i) applying updated versions of FINEMAP and VEP, ii) including SuSiE as a second fine-mapping methodology, providing quasi-replication^11^ to increase the robustness of our findings, particularly for complex regions, iii) analysing complex loci in greater detail by treating distinct credible sets of SNPs individually and iv) exploring less conservative PP thresholds for credible sets. We validated our results by testing the fine-mapped genes against orthogonal signals representing known properties of schizophrenia-associated genes. As in the original PGC study, this included an enrichment analysis for loss of function intolerance (LoFI)^12^. We additionally tested prioritised genes for association between rare coding variant burden and reduced generalised cognition in UK Biobank participants, as recently reported for the original PGC prioritised gene sets^13^.

## Methods

### Fine-mapping

The PGC GWAS meta-analysed a core GWAS, for which individual-level genotypes were available, together with additional datasets where only summary-level data were available. The core GWAS identified 293 linkage disequilibrium independent index SNPs across 255 loci. This core dataset and the associated loci were the basis of the analysis reported here as individual genotype data are required to allow the derivation of in-sample LD for which we used LD-Store (v1.1) (see source GWAS^7^ for further information on summary statistics and LD generation, and region definitions). Fine-mapping was conducted using SuSiE via the susie_rss function of the susieR (0.12.35) package, as well as FINEMAP v1.4.2. The maximum number of causal variants was set to 5 (parameters “L” in SuSiE and “n-causal-snps” in FINEMAP, equivalent to K in the fine-mapping literature) and the sample size was specified as n=161405 (the GWAS sample size).

### SNP/gene annotation

SNPs were annotated to genes using Ensembl VEP^9^ with *Homo sapiens* genome build GRCh37 v115. The default 5000 bp upstream/downstream window was used to annotate SNPs in likely regulatory regions to nearby genes. Where a SNP mapped to multiple Ensembl IDs for the same gene, only the canonical ID was retained. Annotated gene symbols were verified using the HGNChelper R package^14^. Symbols with suggested alternative HGNC-approved names were updated to those (**Supplementary Table 1**).

### Gene prioritisation

The PGC core GWAS was based on participants of European and East Asian ancestry. PGC gene prioritisation was limited to 234 of the 255 core GWAS loci that were replicated in the extended GWAS that also included African and Latino samples, as well as further summary statistics from deCODE genetics. To identify a wider range of genes implicated by schizophrenia genetic architecture, our prioritisation approach was conducted on all 255 fine-mapping loci (notwithstanding exclusion criteria outlined below).

Genes were prioritised if they contained: i) the entire 95% credible set, or ii) a SNP with a PP ≥ 0.1 determined by VEP to be non-synonymous (‘splice_acceptor_variant’/’splice_donor_variant’ or ‘missense_variant’) or to map to a UTR^15^. We next tested prioritisations using credible sets containing 90%, 85%, and 80% of the cumulative PP total.

In the published FINEMAP analysis^7^, gene prioritisation was limited to loci where the best-fitting model suggested the number of causal variants to be three or fewer (k < 3.5). Here, we retained more complex loci, excluding them only if FINEMAP or SuSiE estimated exactly five causal variants (k = 5; FINEMAP: 8 loci excluded, SuSiE: no exclusions). We made this exclusion as such a result is ambiguous and compatible with more independent signals being present at the locus. While these loci could be re-analysed by re-running the software with larger values for the maximum number of causal variants parameter^16^, we considered such complexity most likely to indicate subtle errors or mismatches between GWAS effect sizes and in-sample LD matrices, as previously described^17^.

Note that in the present analysis, we intentionally did not apply functional genomics annotations as our aim was to maximise findings from purely a statistical fine-mapping perspective, an approach which does not rely on integrating data from specific tissues, cells, or developmental stages, and therefore may confer biases based upon preconceptions about the underlying pathophysiology of schizophrenia.

### Loss of function intolerance (LoFI)

LoFI was quantified using the LOEUF (loss-of-function observed/expected upper bound fraction) and pLI (intolerance to heterozygous predicted loss of function variation) metrics from the gnomAD v2.1.1 database^18^. Augmented Mann-Whitney U^19^ (aMWU) was used to assess whether LOEUF and pLI differed between prioritised gene lists and a background gene set, adjusting for standardised gene length. The gene lists were: i) genes prioritised by both FINEMAP and SuSiE; ii) genes prioritised by FINEMAP; iii) genes prioritised by SuSiE and iv) genes prioritised by either FINEMAP or SuSiE but not both. In each case, the background gene set for testing LoFI comparison was the non-prioritised genes mapping to the same locus as a priority gene. For the analysis of the list of genes prioritised by only one of FINEMAP or SuSiE, genes prioritised by both methods at the same loci were excluded from the background gene list. Similarly, for the analysis of the list of additional genes prioritised by relaxing the credible set threshold from 95% to 80% (excluding those prioritised at 95%), those that were prioritised by 95% credible sets were removed from the background gene list,

### Rare coding variants associated with generalised cognition

Some of the present authors have shown that rare deleterious missense variants within genes prioritised by the PGC GWAS of schizophrenia are associated with reduced generalised cognition^20^ using whole-exome sequencing (WES) data from 376,728 unrelated UK Biobank (UKBB) participants of predicted European-like genetic ancestry. Following the methodology of that study, rare coding variants (RCVs) in the UKBB WES data were defined as single nucleotide variants or insertions or deletions <50 bp in length, with minor allele count ≤ 5 in the 376,728 participants. RCVs were classified as protein-truncating (PTV), deleterious missense, or synonymous. PTVs were defined as splice donor, splice acceptor, stop-gain or frameshift variants annotated by VEP, which were also annotated as high confidence for causing loss of protein function by LoFTEE^21^. Deleterious missense variants were annotated as missense by VEP and filtered to those with a REVEL score > 0.75^22^.

Rare synonymous variants, analysed as a negative control, were defined by VEP annotation. ‘*g*’, a measure of generalised cognition, was derived from a principal component analysis of four UKBB cognitive assessments: i) numeric memory; ii) reaction time; iii) pairs matching and iv) alphanumeric trail making test. Measures of *g* were available for 75,188 unrelated UKBB participants of predicted European-like genetic ancestry with WES data. Full details are given in the previous paper^20^.

RCVs in prioritised gene sets were tested for association with *g* in the UKBB. Gene sets tested comprised genes prioritised by i) both FINEMAP and SuSiE by 95% credible sets and/or UTR/non-synonymous variants; ii) both FINEMAP and SuSiE by 80% credible sets and/or UTR/non-synonymous variants; iii) either FINEMAP or SuSiE (but not both) by 95% credible sets and/or UTR/non-synonymous variants; iv) by either FINEMAP or SuSiE (but not both) by 80% credible sets and/or UTR/non-synonymous variants.

### Statistical analysis

Analyses were conducted using R 4.5.1. Z-score standardised gene length was included as a covariate for the aMWU tests. Background gene lists were generated by filtering the gnomAD LoFI dataset for genes that fully or partially intersected with loci boundaries. The aMWU tests yielded marginal probabilistic indices (0.5 = chance probability), and 95% confidence intervals and *p*-values were calculated for the likelihood of prioritised genes having higher LOEUF or pLI than background genes. We used Wald-type interaction tests^23^ to compare LoFI enrichment effect sizes between gene sets, specifically whether genes added by relaxing credible-set thresholds were less enriched than those retained at the most conservative definition (95% credible set and supported by both methods).

A paired Wilcoxon signed rank test was used to test whether the number of independent causal variants inferred at each locus differed between FINEMAP and SuSiE, excluding loci where no credible set was produced by either method.

Linear regression was used to test for association between *g* and RCV burden (deleterious missense, PTV and synonymous) in genes within the prioritised gene sets in the UKBB. Covariates included were exome-wide burden of synonymous variants (except for analyses of synonymous variant burden), sequencing batch, UKBB assessment centre, sex, standardised age, standardised age^2^, sex * standardised age and sex * standardised age^2^, genetic ancestry probabilities and within-ancestry principal components 1-10. Z tests were used to determine if enrichment for RCVs associated with *g* differed between gene sets.

### Synaptic enrichment

The SynGO portal (syngoportal.org)^24^ was used to retrieve synaptic annotations and generate sunburst plots (data retrieved 13/03/26). The default SynGO list of brain-expressed genes was used as the background.

## Results

### Fine-mapping and gene prioritisation

Fine-mapping based on 95% credible sets and UTR and non-synonymous variants (referred to as 95% CS + UTR/NS) prioritised 108 genes using FINEMAP and 97 genes using SuSiE of which 87 were supported by both methods. Fine-mapping based on 80% credible sets, and UTR and non-synonymous variants (referred to as 80% CS + UTR/NS) prioritised 123 genes using FINEMAP and 105 genes using SuSiE, with an overlap of 98 genes (**Figure 1**). Twenty two genes prioritised by FINEMAP and 18 by SuSiE (17 genes in overlap) were based on non-synonymous or UTR SNPs (PP ≥ 0.1). Full FINEMAP results are given in **Supplementary Table 2** and those for prioritised genes are given in **Supplementary Table 3**. Equivalent results for SuSiE are provided in **Supplementary Tables 4** and **5** Of 108 FINEMAP-prioritised genes by 95% CS + UTR/NS, 62 were also prioritised in the PGC analysis^7^, and 46 were novel. Of 97 SuSiE-prioritised genes by 95% CS + UTR/NS, 59 were prioritised in the PGC analysis, and 38 were novel. An additional 9 genes (FINEMAP) and 8 genes (SuSiE) were added by 80% CS + UTR/NS, none in the PGC prioritised list.

**Figure 1.** The distribution of genes across each category of prioritisation. Genes were initially prioritised by either FINEMAP or SuSiE if every SNP within a credible set was annotated to a single gene, or a SNP with a PP ≥ 0.1 was predicted to have a UTR or non-synonymous (missense or splice variants) consequence. Genes were prioritised based on 95%, 90%, 85% and 80% credible sets (80% and 95% credible sets shown here). The upset plot shows the frequency of genes prioritised by each combination of criteria. Set size shows the number of genes prioritised by each broad category.

### Loss of function intolerance

At 95% CS + UTR/NS, all four gene lists showed LoFI enrichment relative to background genes. Lower enrichment (though not significantly different) was seen for gene sets prioritised by only one method (**Figure 2a**; **Supplementary Table 6a**).

**Figure 2.**
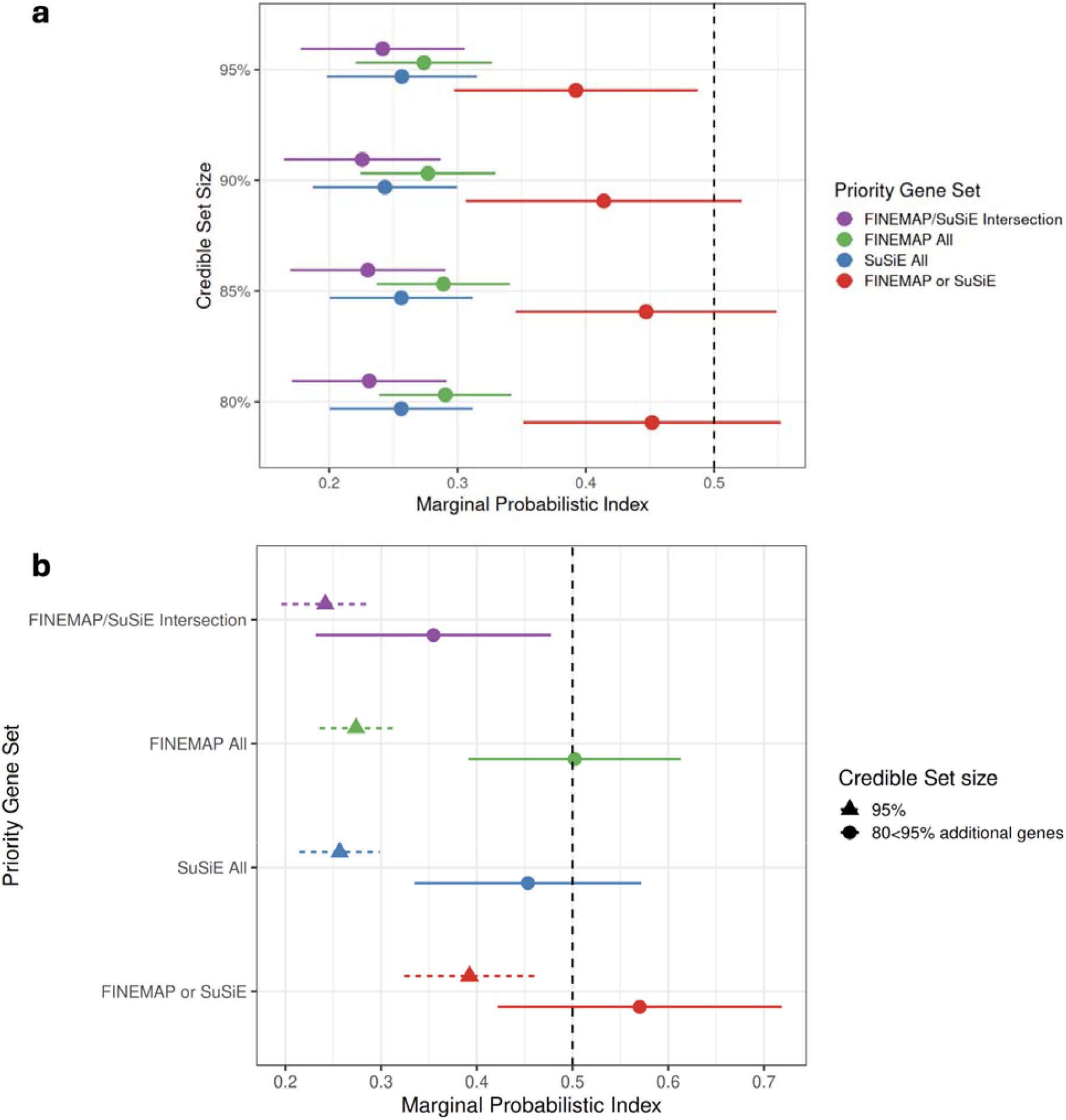
**a** LOEUF score enrichment and 95% confidence intervals for genes prioritised by: both FINEMAP and SuSiE; all genes prioritised by FINEMAP; all genes prioritised by SuSiE and genes prioritised by only one of either FINEMAP or SuSiE. Prioritised gene lists were generated from 95%, 90%, 85% and 80% credible set sizes from FINEMAP and SuSiE + UTR/NS variants. Dashed vertical at marginal probabilistic index (MPI) 0.5 indicates chance level probability of LOEUF enrichment. Lower LOUEF values indicate greater LoFI and therefore lower MPI indicates higher LoFI enrichment. **b** Comparison of LoFI (LOEUF) enrichment effect sizes between 95% CS + UTR/NS prioritised genes and additional genes prioritised by loosening credible set size to 80% + UTR/NS for both FINEMAP and SuSiE intersecting genes, all FINEMAP, all SuSiE, and genes prioritised by only FINEMAP or SuSiE. Effect sizes in panel a are visualised with 95% confidence intervals, whereas effect sizes in panel b are visualised with 84% confidence intervals to allow visual estimation of significant difference between sets rather than to the null hypothesis.

Relaxing the requirement that the full 95% credible set was contained within a single gene to the sum of the PP of at least 90%, 85%, and 80% (+ UTR/NS variants), genes prioritised by the intersection of methods, as well as all FINEMAP and all SuSiE genes were enriched for LoFI. Those prioritised by only by FINEMAP or by SuSiE were not significantly enriched (**Figure 2a**).

Reducing the credible set size from 95% + UTR/NS to 80% + UTR/NS identified an additional 11 prioritised genes supported by both FINEMAP and SuSiE. Although underpowered for a separate analysis, these 11 additional genes had at a trend level lower average LOEUF values compared to background genes at the same loci (P = 0.096). Genes prioritised by relaxing the credible set criterion to 80% that were only prioritised by one fine-mapping method were not enriched for LoFI compared with other genes at the same loci (P = 0.505) (**Figure 2b**). Full results including those based on pLI can be found in **Supplementary Table 6b**.

Genes supported only at 80% CS + UTR/NS (and not at 95% CS + UTR/NS) did not significantly differ in LOEUF enrichment from those that were supported by both FINEMAP and SuSiE at the more conservative 95% CS (P = 0.225; **Figure 2b**). In contrast, genes prioritised at 95% CS by only one method were significantly less enriched for LOEUF compared to genes supported by both methods at that threshold (P = 0.010), and so were additional genes prioritised at 80% CS by only one method (P = 0.003). Full results including pLI can be found in **Supplementary Table 6c**.

### Rare coding variants associated with generalised cognition

A higher burden of deleterious missense variants in genes prioritised at 95% CS + UTR/NS by both FINEMAP and SuSiE was associated with lower *g* (P = 4.57x10^-6^); a higher burden in genes prioritised at 80% CS + UTR/NS by both methods was similarly associated (P = 5.17x10^-6^; **Figure 3**). This was not the case for genes prioritised by only one fine-mapping method at either 95% (P = 0.855) or 80% CS sizes (P = 0.894). No associations were found for rare PTVs or synonymous RCVs with any gene sets (full results reported in **Supplementary Table 7**).

**Figure 3.**
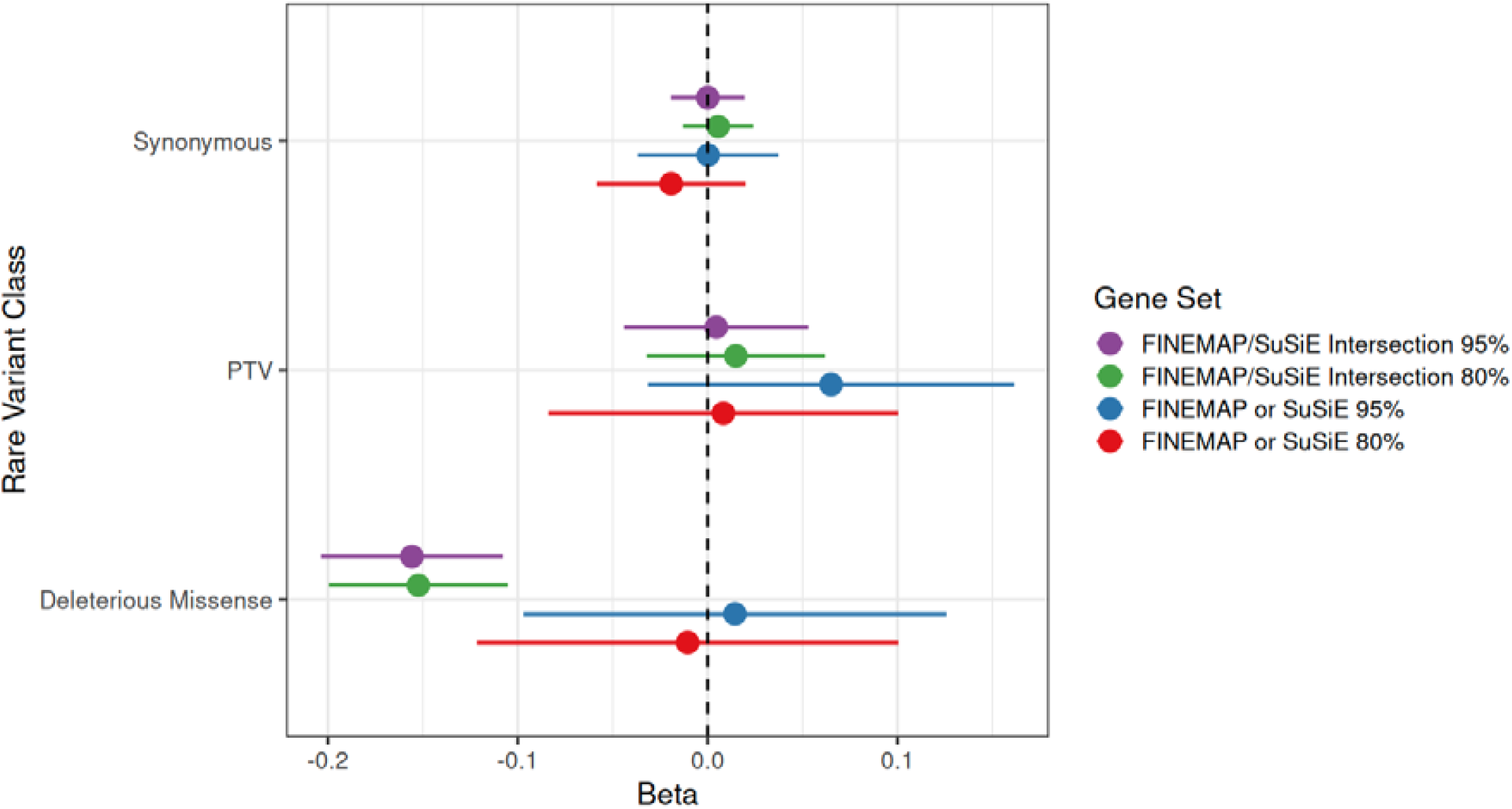
Association between RCV burden (deleterious missense, PTV and synonymous) and *g* in UKBB participants in gene sets prioritised by i) 95% CS + UTR/NS in both FINEMAP and SuSiE; ii) 80% CS + UTR/NS in both FINEMAP and SuSiE, iii) 95% CS + UTR/NS in FINEMAP or SuSiE, but not both; iv) 80% CS + UTR/NS in FINEMAP or SuSiE, but not both. Effect sizes are visualised with 84% confidence intervals to allow visual estimation of significant difference between sets rather than to the null hypothesis.

Association between missense burden and *g* was greater in 95% CS + UTR/NS genes prioritised by both FINEMAP and SuSiE compared with 95% CS + UTR/NS genes prioritised by only one method (*Z* = -1.98, P = 0.048) (**Figure 3**).

### Final Prioritisation

Guided by the LoFI and the cognition analyses, we defined highest confidence prioritised genes (N=98, 89 protein-coding) as those which contained the sum of at least 80% of the PP in both FINEMAP and SuSiE, and/or genes in which a SNP with a PP ≥ 0.1 mapped to a UTR/NS effect (**Figure 4**). A full list of genes prioritised by FINEMAP and/or SuSiE is included in **Supplementary Table 8a**.

**Figure 4.**
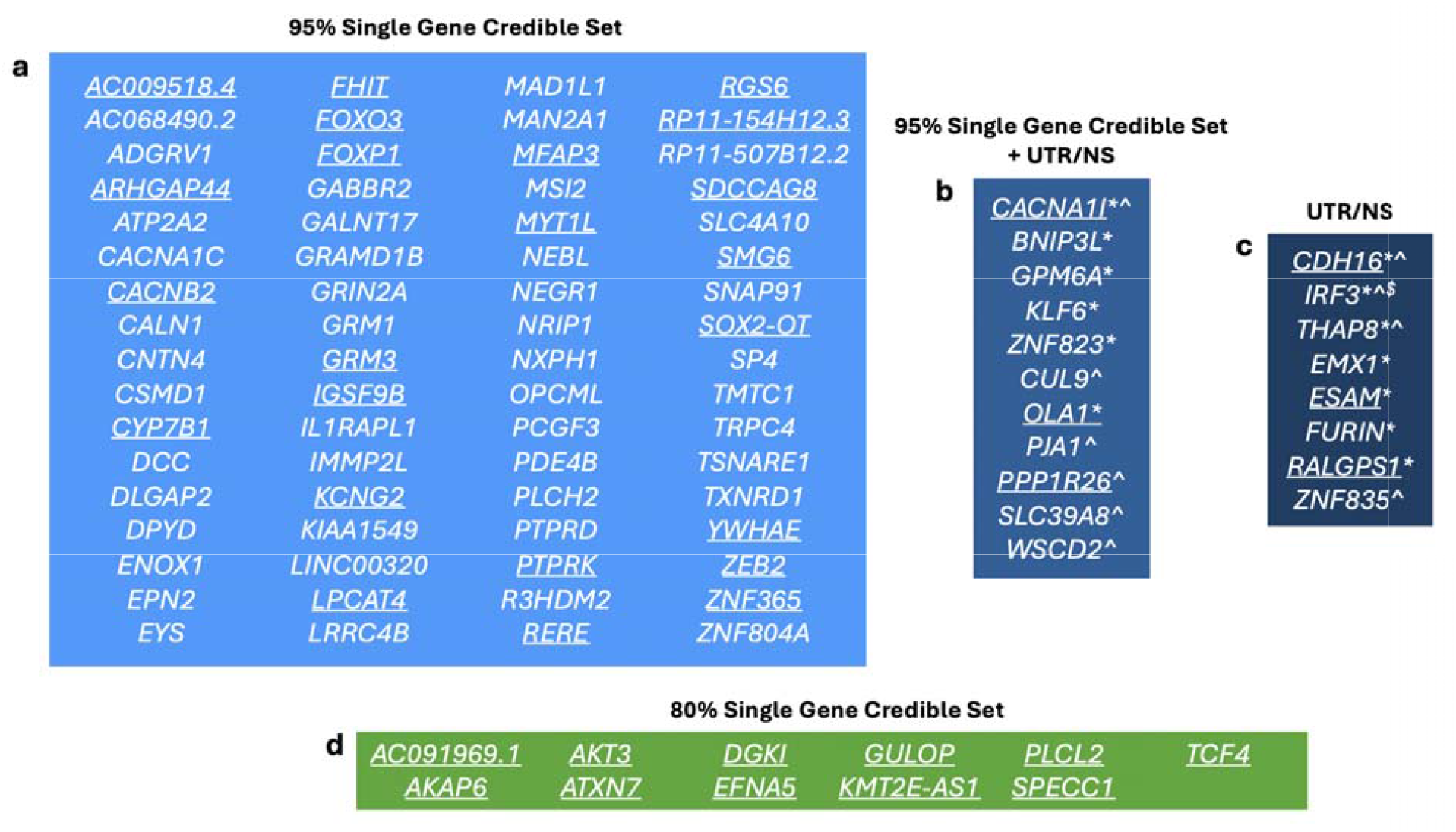
Final prioritised genes. **a** 95% single gene credible set genes were prioritised by both FINEMAP and SuSiE when all SNPs within a 95% credible were annotated to that gene. **b** UTR/NS genes were prioritised by both 95% single gene credible sets (as in a) and when a SNP with PP ≥ 0.1 was predicted by VEP to have a UTR or non-synonymous (splicing or missense) effect (as in c) in FINEMAP and SuSiE. **c** Genes prioritised by both FINEMAP and SuSiE by UTR/NS but not by a gene annotating to an entire 95% credible set. **d** 80% single gene credible set genes were prioritised by both FINEMAP and SuSiE when credible set sizes were relaxed from 95% to 80% when all SNPs within a credible set were annotated to that gene. Underlined genes were not prioritised in the 2022 PGC analysis (n = 40) based on statistical fine-mapping alone although several (e.g. *CYP7B1, AKT3, RERE*) were prioritised based on functional genomic data while others (*FHIT, FOXP1, LPCAT4, MYTL1, PTPRK, CDH16, PPP1R26*) would have met fine-mapping prioritisation criteria but were excluded as loci were not significant in the extended GWAS. ADGRV1 and GALNT17 were prioritised in the PGC analysis but have undergone name changes from GPR98 and WBSCR17 respectively. * predicted UTR effect; ^ predicted missense effect; ^$^ predicted splicing effect.

Twelve PGC FINEMAP genes^7^ were not replicated by this new prioritisation (**Supplementary Table 8b**). Four (*ACTR1B, BCL11B, NLGN4X*, and *SGCD)* were, as in the original analysis, supported by FINEMAP, but not by SuSiE. Three (*BCL2L12, C12orf43* and *PCNX3*) could not be prioritised due to changes in overlap between credible sets and genes. Finally, five (*CTD-2008L17*.*2, LINC01088, NAB2, PAK6* and *RP11-399D6*.*2*) were excluded because their 95% CS credible SNPs mapped to more than one gene simultaneously, impairing a clear prioritisation. The PGC analysis did not have these ambiguous multi-gene mappings as only the gene annotated with the most severe VEP consequence was retained, while our current use of VEP preserved all predicted consequences and annotations for parsimony.

### Synaptic Enrichment

Of the 89 protein-coding genes prioritised by both FINEMAP and SuSiE 80% CS + UTR/NS, 23 (25.8%) were annotated to synaptic functioning according to the SynGO database (**Figure 5**).

**Figure 5.**
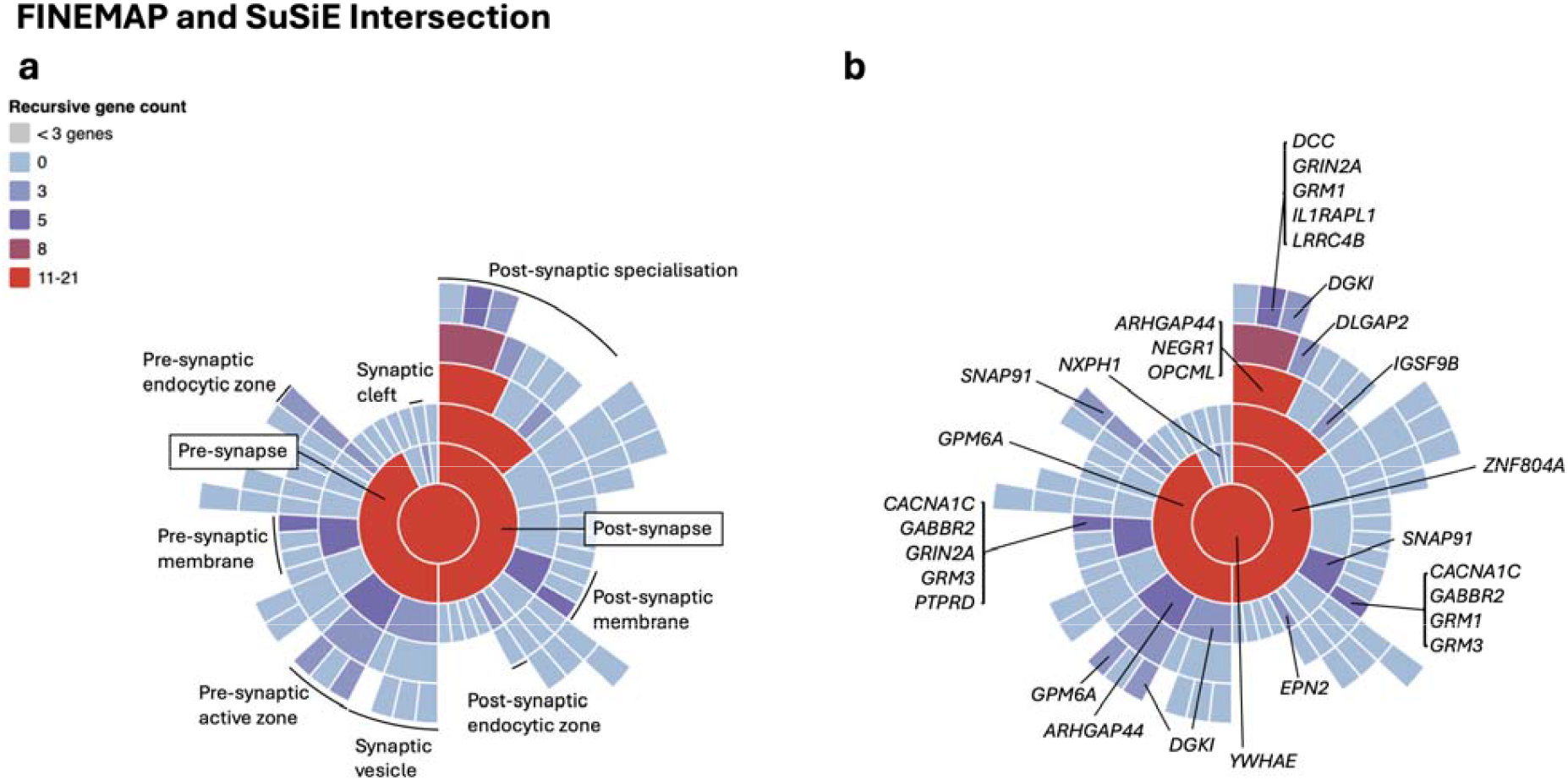
Sunburst plots depicting synaptic locations starting with the synapse (centre), pre- and postsynaptic locations in the first ring and child terms in outer rings. The legend indicates the number of genes corresponding to each term. **a** and **b**, protein-coding genes prioritised by both FINEMAP and SuSiE at 80% CS + UTR/NS (n = 89).

Genes prioritised by both FINEMAP and SuSiE were enriched for six gene ontology (GO) biological process terms after FDR p-value correction (significance threshold: P_FDR_ = 0.05): ‘trans-synaptic signaling’ (P = 1.30x10^-4^); ‘process in the synapse’ (P = 1.45x10^-4^); ‘chemical synaptic transmission’ (P = 1.45x10^-4^); ‘synapse adhesion between pre- and post-synapse’ (P = 2.34x10^-4^); ‘modulation of chemical synaptic transmission’ (P = 2.64x10^-3^) and ‘synapse organization’ (P = 2.64x10^-3^). The gene list was also enriched for 11 cellular component terms, spanning pre- and post-synaptic compartments: ‘postsynapse’ (P = 6.81x10^-5^); ‘postsynaptic specialisation’ (P = 6.81x10^-5^); ‘postsynaptic density’ (P = 6.81x10^-5^); ‘synapse’ (P = 1.35x10^-4^); ‘postsynaptic density membrane’ (P = 2.39x10^-4^); ‘integral component of postsynaptic density membrane’ (P = 6.30x10^-4^); ‘integral component of presynaptic membrane’ (P = 9.45x10^-4^); ‘postsynaptic membrane’ (P = 1.65x10^-3^); ‘integral component of postsynaptic membrane’ (P = 4.28x10^-3^); ‘presynaptic active zone’ (P = 1.94x10^-2^) and ‘presynapse’ (P = 2.29x10^-2^).

### Causal variant structure

Eight loci where FINEMAP estimated exactly five causal variants were removed from gene prioritisation (**Supplementary Table 1b**). For both FINEMAP and SuSiE, the majority of loci were predicted to have a single causal variant (FINEMAP n = 163; SuSiE n = 193).

FINEMAP’s range of estimated causal variants (k=1-5) exceeded SuSiE (k=1-3), FINEMAP tending to infer more causal variants per locus than SuSiE (*Z* = 6.275, *W* = 1378, P = 5.15x10^-11^).

### Complex locus CHR18:52716708-53630258 (*TCF4*)

*TCF4* was prioritised by FINEMAP at 95% CS and SuSiE at 80% CS. This locus spans four merged LD clumps (index SNPs rs12969453, rs74914300, rs72934602 and rs9636107) and was predicted to have four independent causal variants by FINEMAP and the PGC conditional analysis^7^, while SuSiE estimated three (**Figure 6**).

**Figure 6.**
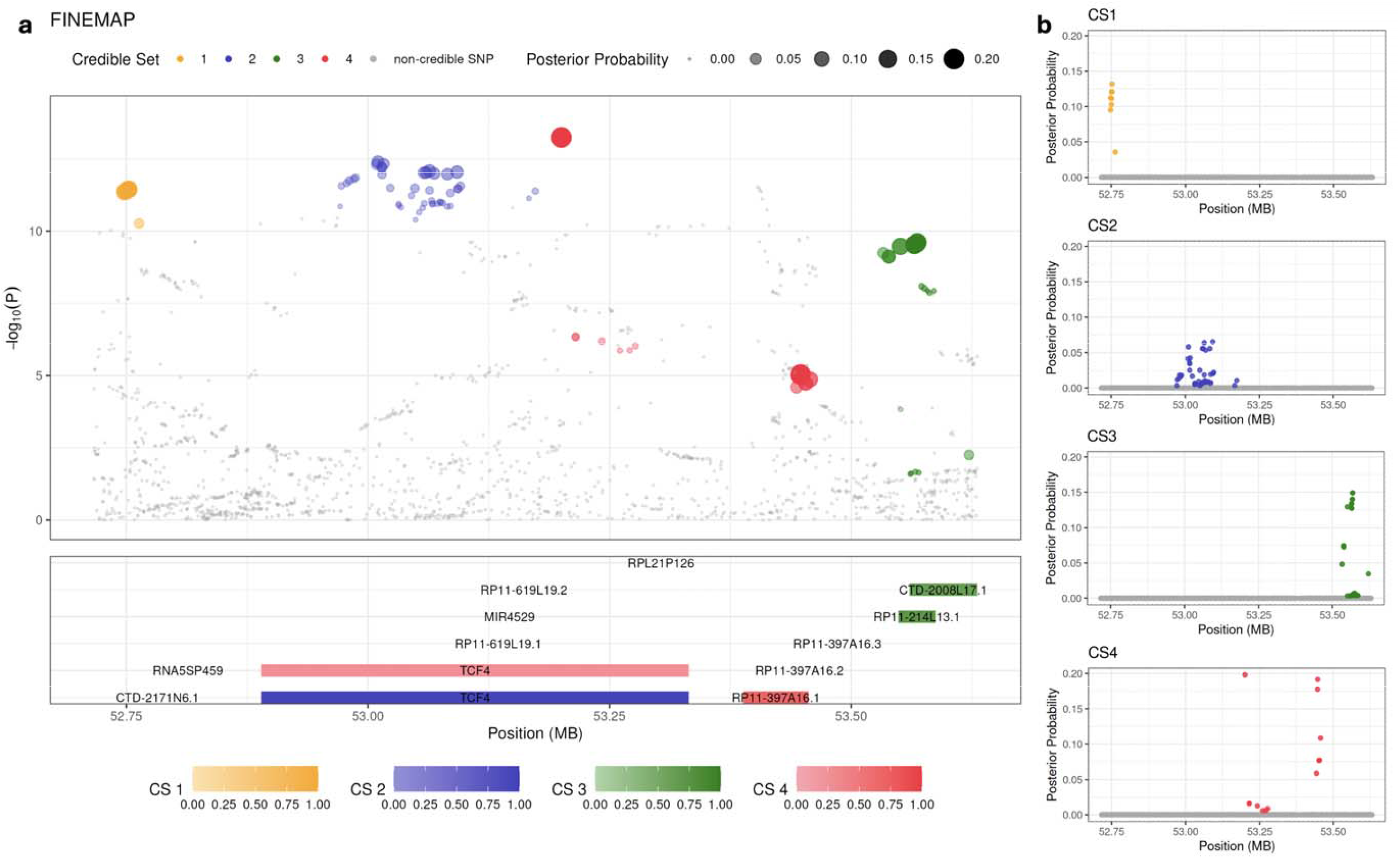
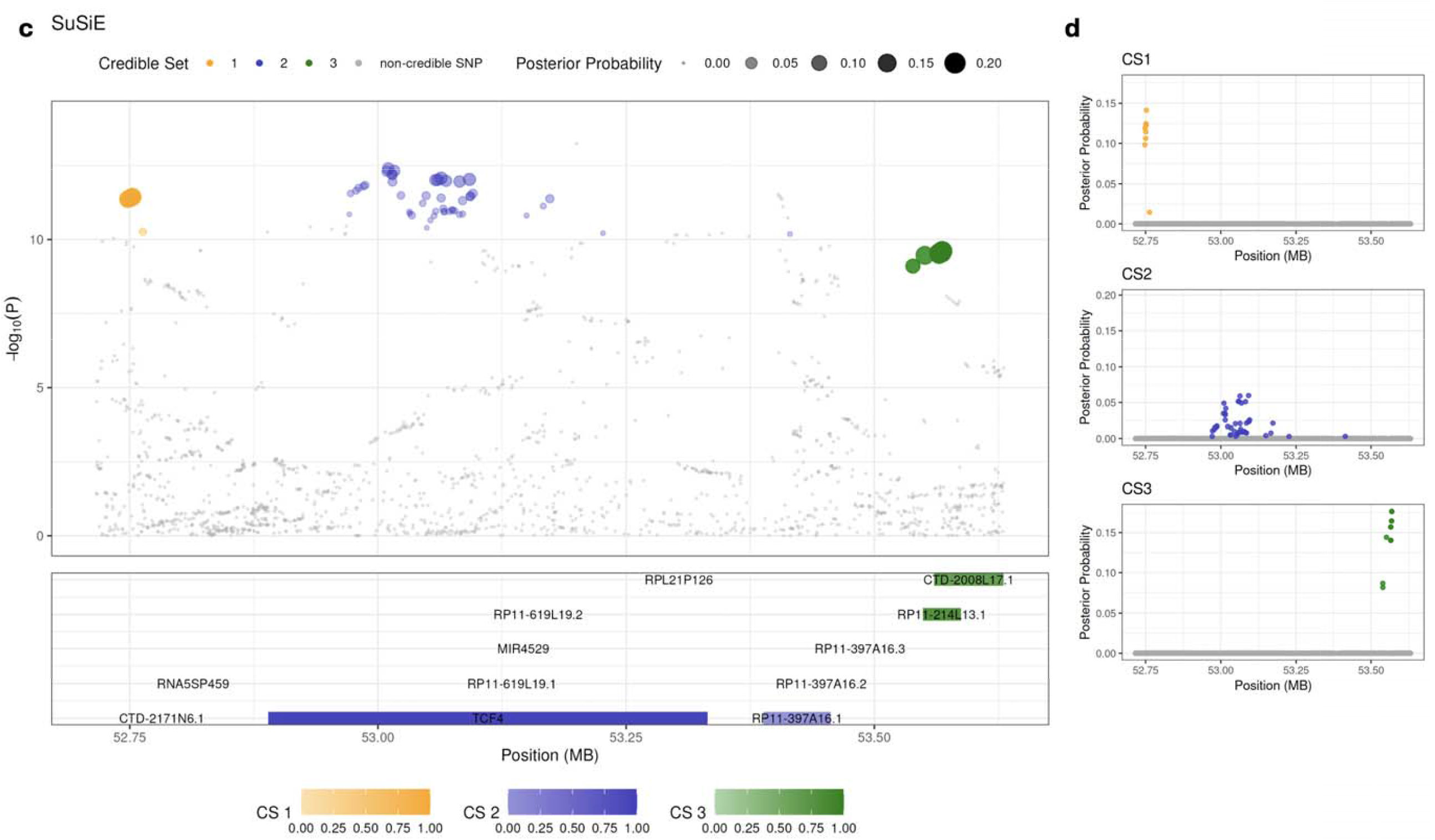
Locus CHR18:52716708-53630258. **a** and **b**: FINEMAP. **c** and **d**: SuSiE. **a** and **c**: top panel: -log_10_(P) of each SNP at the locus in the PGC schizophrenia GWAS against the base position on chromosome 18. Colour of SNPs refer to whether they are contained within a 95% credible set, and size indicates PP. Bottom panel: gradient colour of genes refers to the 95% credible set in which they are implicated and the cumulative PP of SNPs annotated to that gene. **b** and **d**: PP for each SNP within the locus against base position (MB). SNP colour denotes presence within the 95% credible set.

*TCF4* was prioritised by FINEMAP CS2 containing 44 SNPs, all annotated to *TCF4*. All 44 SNPs are also contained within SuSiE CS2 which contains 47 SNPs, of which all but 1 (with a negligible PP of only 0.003) annotate to *TCF4*. FINEMAP and SuSiE CS1 (see top panels **Figure 6 a,c**) are concordant, both containing the same nine SNPs, none annotated to any gene. FINEMAP and SuSiE CS3 are also highly concordant: the SuSiE credible set contains seven SNPs, all included in the FINEMAP credible set, but the latter contains an additional 12 SNPs (the additional SNPs with a total PP of 0.126). Both FINEMAP and SuSiE CS3s contain SNPs annotated to CTD-2008L17.1 and RP11-214L13.1; neither gene was prioritised because of ambiguous multi-gene mapping.

## Discussion

By applying two independent Bayesian statistical fine-mapping methods (FINEMAP and SuSiE) to the 2022 PGC schizophrenia GWAS, extending the analysis to complex loci with multiple causal variants, and relaxing credible set thresholds, we identified 98 high-confidence schizophrenia priority genes, including 40 not named in the original study. Genes supported by both FINEMAP and SuSiE were strongly enriched for LoFI metrics^12^, and were enriched for burden of rare deleterious missense mutations associated with reduced generalised cognition^20^. These associations are established features of schizophrenia-associated gene sets, providing orthogonal validations for our prioritisation procedure.

The support of genes by both FINEMAP and SuSiE had important implications for prioritisation. Those prioritised at 95% CS + UTR/NS were enriched for LoFI metrics irrespective of whether one or both methods supported them, but nevertheless, those prioritised by only one method were significantly less enriched for LoFI than those supported by both. The same pattern was seen in the cognition analyses: RCV burden in genes prioritised by both methods was associated with reduced *g* in UKBB participants, but genes prioritised by only one method were not. The LoFI enrichment analysis results suggests that biologically relevant genes may still be present amongst those supported by only one fine-mapping method, but the overall attenuation in both the validation analyses reduces the degree of confidence in this set of genes as a whole. Aiming to be conservative, we focus only on genes supported by both FINEMAP and SuSiE in our final prioritised gene list.

Genes prioritised by both methods at a relaxed credible set threshold (cumulative PP ≥ 80%) were still enriched for LoFI, and for missense burden associated with reduced generalised cognition. The level of LoFI enrichment of the additional genes prioritised by both methods when relaxing the credible set threshold from 95% to 80% did not differ from those prioritised at a 95% level, although due to the small gene set size (N=11), it was not possible to reliably test the association between RCV burden and generalised cognition in these additional genes alone. Genes prioritised by only one method at the 80% credible set threshold did not pass either validation test. Thus, our final list of prioritised genes included those prioritised by both FINEMAP and SuSiE at the 80% credible set size, as well as those with predicted UTR or non-synonymous effects. The combined use of these two fine-mapping methods allows for a more conservative approach to the identification of schizophrenia high priority genes, as validated by LoFI enrichment and deleterious missense variant burden association with reduced generalised cognition.

Of the final 98 prioritised genes, 40 were not prioritised in the PGC publication (**Supplementary Table 5a**). Consistent with current understanding of schizophrenia biology^7,25^, prioritised genes were enriched for synaptic biology and included seven additional genes that were not previously prioritised (*ARHGAP44, CACNB2, DGKI, EFNA5, GRM3, IGSF9B, YWHAE*) annotated as having synaptic functions. Several of the prioritised genes show minimal expression in brain in GTEx v8^26^ (*CDH16, EYS, GULOP, IL1RAPL1* and *MAD1L1*), with *CDH16* showing highly specific expression in renal and thyroid tissues. Thus, some of the common variant genetic architecture of schizophrenia may increase risk via aetiological pathways outside of the brain, although some are likely to represent false positives as expected from prioritisation approaches.

By treating individual credible sets at complex loci independently, we prioritised 15 novel genes (*AC009518*.*4, AKT3, ARHGAP44, CACNB2, CYP7B1, GRM3, IGSF9B, KCNG2, MFAP3, RGS6, RP11-154H12*.*3, SDCCAG8, YWHAE, ZEB2* and *ZNF365*), several of which have established roles in synaptic function and neurodevelopment. Six further novel genes reflect changes in the FINEMAP model since the initial analysis: *AKAP6, CACNA1I, ESAM, FOXO3, OLA1* and *RERE* while updated VEP SNP annotations prioritised *RALGPS1*, SMG6, and SOX2-OT. A further seven genes (*CDH16, FHIT, FOXP1, LPCAT4, MYT1L, PPP1R26* and *PTPRK*) were prioritised from loci that were excluded from the PGC gene prioritisation because they were not replicated in the extended GWAS. Further work with larger non-European samples is required to determine whether any ancestry-specific effects are underlying these associations.

FINEMAP tended to predict more causal variants per locus than SuSiE. This systematic difference likely reflects SuSiE’s ‘*purity’* filter (see **Supplementary Note** for more detail), rather than genuine disagreement across methods about which loci harbour a complex architecture. Indeed, where both methods produced multiple credible sets, concordance was high. We provide the complex locus surrounding the *TCF4* gene at 18q21.2 by way of illustration. *TCF4* plays a crucial role in neurodevelopment^27^. The locus was implicated in schizophrenia in one of the first schizophrenia GWAS studies, which also supported the presence of multiple independent causal signals at the locus^28^. In the present analysis, FINEMAP produced four independent credible sets and SuSiE produced three, with most causal variants and credible set locations overlapping (**Figure 6**). *TCF4* was prioritised by both methods, adding further support for the importance of this gene in schizophrenia aetiology. Whether the independent signals at this locus act through *TCF4* regulatory function or via independent mechanisms remains unknown.

Variants with non-synonymous (missense/splicing) and UTR effects are enriched for causal SNPs across complex traits^29^. The present study included 19 genes with possible causal UTR/no-synonymous variants, of which 11 were also prioritised based on credible set size.

Six of these were not prioritised in the PGC analysis: *CACNA1I, CDH16, ESAM, OLA1, PPP1R26* and *RALGPS1. PPP1R26* was prioritised with considerably high support, by a single high-confidence SNP (PP = 0.99; inferred by FINEMAP and SuSiE) with a predicted missense effect, and by the nature of a being a credible set comprising a single SNP, it was also prioritised as a 95% single gene credible set. *CDH16* and *RALGPS1* were also well supported, and were prioritised by SNPs with moderate cumulative PPs (≥0.3; both methods), both with predicted UTR effects, and *CDH16* additionally prioritised by a SNP with a predicted missense effect.

We were unable to prioritise genes for 178 (69.8%) loci. In most cases, this reflected a large number of genes tagged by the credible set SNPs, often with overlapping boundaries. A dearth of tagged genes was a rarer occurrence, and nine loci had contained no annotated at all. Resolving these loci is likely to require approaches that incorporate functional annotations, such as eQTLs or chromatin loops, to identify which genes are regulated by the credible causal variants.

In summary, we have substantially advanced earlier attempts to fine-map schizophrenia loci, identifying 98 highly credible causal genes. Of these, 40 have not been previously prioritised by statistical fine-mapping, providing a substantially extended set of targets for downstream functional follow-up aiming to determine the molecular mechanisms through which genes contribute to the aetiology of schizophrenia.

## Data Availability

All data produced in the study are contained in the manuscript, supplementary files or available upon reasonable request to the authors. This research was also conducted using the UK Biobank Resource under Application Number 13310. Data from the UK Biobank is available for health-related research upon registration and application through the UK Biobank Access Management System.
All data used in the manuscript are available online at the following websites.

https://pgc.unc.edu/for-researchers/data-access-committee/data-access-portal/

https://pgc.unc.edu/for-researchers/download-results/

https://www.ukbiobank.ac.uk/enable-your-research/register

## References

1. Vigo, D., Thornicroft, G. & Atun, R. Estimating the true global burden of mental illness. Lancet Psychiatry 3, 171–178 (2016).

2. Bienvenu, O. J., Davydow, D. S. & Kendler, K. S. Psychiatric ‘diseases’ versus behavioral disorders and degree of genetic influence. Psychol. Med. 41, 33–40 (2011).

3. Hilker, R. et al. Heritability of Schizophrenia and Schizophrenia Spectrum Based on the Nationwide Danish Twin Register. Biol. Psychiatry 83, 492–498 (2018).

4. Smeland, O. B., Frei, O., Dale, A. M. & Andreassen, O. A. The polygenic architecture of schizophrenia — rethinking pathogenesis and nosology. Nat. Rev. Neurol. 16, 366–379 (2020).

5. Owen, M. J., Craddock, N. & O’Donovan, M. C. Suggestion of Roles for Both Common and Rare Risk Variants in Genome-wide Studies of Schizophrenia. Arch. Gen. Psychiatry 67, 667 (2010).

6. Schaid, D. J., Chen, W. & Larson, N. B. From genome-wide associations to candidate causal variants by statistical fine-mapping. Nat. Rev. Genet. 19, 491–504 (2018).

7. Trubetskoy, V. et al. Mapping genomic loci implicates genes and synaptic biology in schizophrenia. Nature 604, 502–508 (2022).

8. Benner, C. et al. FINEMAP: efficient variable selection using summary data from genome-wide association studies. Bioinformatics 32, 1493–1501 (2016).

9. McLaren, W. et al. The Ensembl Variant Effect Predictor. Genome Biol. 17, 122 (2016).

10. Zou, Y., Carbonetto, P., Wang, G. & Stephens, M. Fine-mapping from summary data with the “Sum of Single Effects” model. PLoS Genet. 18, e1010299 (2022).

11. Bettis, R. A., Helfat, C. E. & Shaver, J. M. The necessity, logic, and forms of replication. Strategic Management Journal 37, 2193–2203 (2016).

12. Pardiñas, A. F. et al. Common schizophrenia alleles are enriched in mutation-intolerant genes and in regions under strong background selection. Nat. Genet. 50, 381–389 (2018).

13. Fenner, E. et al. Analysis of rare coding variants in schizophrenia-associated genes and generalised cognition in the UK Biobank. Mol. Psychiatry https://doi.org/10.1038/s41380-026-03601-8 (2026) doi:10.1038/s41380-026-03601-8.

14. Oh, S. et al. HGNChelper: identification and correction of invalid gene symbols for human and mouse. F1000Res. 9, 1493 (2020).

15. EMBL-EBI. Ensembl Variation - Calculated variant consequences. https://www.ensembl.org/info/genome/variation/prediction/predicted_data.html (2025).

16. Li, Z. & Zhou, X. Towards improved fine-mapping of candidate causal variants. Nat. Rev. Genet. 26, 847–861 (2025).

17. Benner, C. et al. Prospects of Fine-Mapping Trait-Associated Genomic Regions by Using Summary Statistics from Genome-wide Association Studies. The American Journal of Human Genetics 101, 539–551 (2017).

18. Karczewski, K. J. et al. The mutational constraint spectrum quantified from variation in 141,456 humans. Nature 581, 434–443 (2020).

19. Vermeulen, K., Thas, O. & Vansteelandt, S. Increasing the power of the Mann-Whitney test in randomized experiments through flexible covariate adjustment. Stat. Med. 34, 1012–1030 (2015).

20. Fenner, E. et al. Rare Coding Variants in Schizophrenia-Associated Genes Affect Generalised Cognition in the UK Biobank. (2025) doi:10.1101/2023.08.14.23294074.

21. Karczewski, K. J. et al. The mutational constraint spectrum quantified from variation in 141,456 humans. Nature 581, 434–443 (2020).

22. Ioannidis, N. M. et al. REVEL: An Ensemble Method for Predicting the Pathogenicity of Rare Missense Variants. The American Journal of Human Genetics 99, 877–885 (2016).

23. Altman, D. G. Statistics Notes: Interaction revisited: the difference between two estimates. BMJ 326, 219–219 (2003).

24. Koopmans, F. et al. SynGO: An Evidence-Based, Expert-Curated Knowledge Base for the Synapse. Neuron 103, 217–234.e4 (2019).

25. Hall, J., Trent, S., Thomas, K. L., O’Donovan, M. C. & Owen, M. J. Genetic Risk for Schizophrenia: Convergence on Synaptic Pathways Involved in Plasticity. Biol. Psychiatry 77, 52–58 (2015).

26. Aguet, F. et al. The GTEx Consortium atlas of genetic regulatory effects across human tissues. Science (1979). 369, 1318–1330 (2020).

27. Li, H. et al. Disruption of TCF4 regulatory networks leads to abnormal cortical development and mental disabilities. Mol. Psychiatry 24, 1235–1246 (2019).

28. Steinberg, S. et al. Common variants at VRK2 and TCF4 conferring risk of schizophrenia. Hum. Mol. Genet. 20, 4076–4081 (2011).

29. MacLeod, I. M. et al. Exploiting biological priors and sequence variants enhances QTL discovery and genomic prediction of complex traits. BMC Genomics 17, 144 (2016).

