## Supplementary Note for "Statistical fine-mapping of schizophrenia common risk loci using FINEMAP and SuSiE"

FINEMAP^1^ and SuSiE^2,3^ are Bayesian statistical fine-mapping methods that do not require access to individual-level genotype data. Their inputs are GWAS summary statistics and locus-wide linkage disequilibrium (LD) matrices, either derived from in-sample genotypes or from a reference panel. Both methods produce a posterior probability for each SNP within a locus, which can be interpreted as the probability of the SNP being the causal variant responsible for the regional GWAS signal. Credible sets of SNPs are constructed by ranking SNPs from highest to lowest posterior and adding them sequentially until their cumulative probability reaches a pre-specified threshold, commonly 95%. A 95% credible set therefore has a 95% probability of containing the true causal SNP, conditional on several assumptions: that the LD matrix is correctly specified, that the trait is influenced by additive SNP effects at the locus, and that the true causal variant is captured by the genotyped or imputed data. While this interpretation applies to the output of both FINEMAP and SuSiE, each algorithm differs in how posterior probabilities are computed. The remainder of this Supplementary Note provides an overview of key algorithmic differences most relevant to potential cross-method discrepancies that may be encountered in practical applications.

At any locus with moderate numbers of SNPs, the number of variants and combinations of variants that may explain the GWAS signal grows rapidly when multiple causal variants are assumed. This makes an exhaustive evaluation computationally impractical. FINEMAP addresses this with a stochastic search strategy (the “shotgun” algorithm^4^): rather than evaluating all possible variant combinations, it probabilistically samples markers and concentrates computation on those most strongly supported as causal by the data, which are termed “SNP configurations”. SuSiE takes a fundamentally different approach based on variational inference^5^: Instead of searching across combinations of variants, its Bayesian stepwise selection works on one marker at a time, assuming a signal caused by multiple causal variants will manifest as a ‘sum’ of variant effects derived from single-effect regression. In both methods, the maximum number of causal effects that may explain the GWAS signal, and that will be evaluated by the software, is user-specified (*K* in FINEMAP, *L* in SuSiE).

The main difference between FINEMAP and SuSiE is the determination of the number of independent causal variants (*k*) at a locus. FINEMAP models *k* explicitly using prior probabilities that decrease as *K* increases, reflecting a parsimonious assumption in which simpler causal structures are always more likely. In this process, SNP configurations are sequentially extracted for each k ∈ {1, 2, …, K}, and posterior probabilities are computed for each configuration. At each step, the probability for that value of *k* being the true value can be computed as the sum of all configurations of posterior probabilities, though this is only an approximation as not all possible SNP combinations are formally evaluated. All the causal models evaluated by FINEMAP can be used to compute the expected value of *k* at each locus, defined as the weighted sum:

$$E[k]=\sum_{k=1}^{K} k\cdot p(k)$$

Critically, this FINEMAP estimate of *k* is determined independently from the assignment of SNPs to credible sets. This is not the case in SuSiE, where *k* is not modelled or evaluated directly. Instead, it is inferred from the number of credible sets retained by the algorithm after a final filtering step influenced by the “purity” metric, which is computed for each credible set as the minimum pairwise correlation between the SNPs included within. This filter reflects that markers within a genuine credible set all represent the same independent causal signal and should therefore be in high linkage disequilibrium with one another. Credible sets that do not meet a certain purity threshold (default *r*=0.5) are more likely to reflect noise and uncertainty than a well-resolved signal and are consequently discarded. The elimination of sets with poor purity reduces SuSiE's inferred *k*, which makes the filter parsimonious in a similar way to FINEMAP’s prior on this value. However, it also introduces a systematic difference between methods, which explains our empirical observations of SuSiE reporting lower k values than FINEMAP and not vice versa, despite the same input data being used in all analyses.

There are subtle differences between the posterior probabilities of SNPs reported by FINEMAP and SuSiE, which are termed PP by FINEMAP and alpha by SuSiE. Because FINEMAP evaluates SNP configurations at each value of *k* (from 1 to *K*), each step in this computation produces its own SNP PPs and credible sets. In contrast, SuSiE returns only the credible sets and alpha values produced by its final post-purity solution. In this work, the credible sets reported in the FINEMAP analysis correspond to those of the highest probability value of *k*, and the PPs are extracted from those. Our use of “posterior probability" in this manuscript refers to FINEMAP PPs and SuSiE alphas, as these are conceptually analogous quantities reflecting the probability of a SNP being causal within a specific causal variant model or single-effect regression component. For clarity, we note the related term of “posterior inclusion probability” (PIP), though we do not use this metric in the prioritisation analyses. In FINEMAP, PIP is the sum of all causal configurations PPs that include that SNP, while in SuSiE, it is the marginal probability across all single-effect regression components. PIPs from both methods can be interpreted in the same way, as locus-wide marginal probabilities of causality.

**References**

1.	Benner, C. *et al.* FINEMAP: efficient variable selection using summary data from genome-wide association studies. *Bioinformatics* **32**, 1493–1501 (2016).

2.	Zou, Y., Carbonetto, P., Wang, G. & Stephens, M. Fine-mapping from summary data with the “Sum of Single Effects” model. *PLoS Genet.* **18**, e1010299 (2022).

3.	Wang, G., Sarkar, A., Carbonetto, P. & Stephens, M. A Simple New Approach to Variable Selection in Regression, with Application to Genetic Fine Mapping. *J. R. Stat. Soc. Series B Stat. Methodol.* **82**, 1273–1300 (2020).

4.	Hans, C., Dobra, A. & West, M. Shotgun Stochastic Search for “Large *p* ” Regression. *J. Am. Stat. Assoc.* **102**, 507–516 (2007).

5.	Blei, D. M., Kucukelbir, A. & McAuliffe, J. D. Variational Inference: A Review for Statisticians. *J. Am. Stat. Assoc.* **112**, 859–877 (2017).
